# Examining the bidirectional association between emotion recognition and autistic traits using observational and genetic analyses

**DOI:** 10.1101/2020.05.21.20108761

**Authors:** Zoe E. Reed, Liam Mahedy, Abigail Jackson, George Davey Smith, Ian Penton-Voak, Angela S Attwood, Marcus R Munafò

## Abstract

**Background:** There is mixed evidence for an association between autism spectrum disorder (ASD) and emotion recognition deficits. We sought to assess the bidirectionality of this association using phenotypic and genetic data in a large community sample.

**Methods:** Analyses were conducted in three stages. First, we examined the bidirectional association between autistic traits at age 8 years and emotion recognition task (ERT) responses at age 24 years (Study 1; N=3,562); and between Diagnostic Analysis of Non-Verbal Accuracy (DANVA) emotion recognition responses at age 8 years and autistic traits at age 10 years (Study 2; N=9,071). Next, we used genetic analyses (Study 3) to examine the association between polygenic risk scores for ASD and these phenotypes. The genetic correlation between ASD and ERT responses at age 24 was also estimated. Analyses were conducted in the Avon Longitudinal Study of Parents and Children.

**Results:** Autistic traits at age 8 years were negatively associated with later total correct responses on ERT in Study 1 *(b=-*0.18; 95% CI: −0.27 to −0.09). We also found evidence of an association in Study 2 (*b*=-0.04; 95% CI: −0.05 to −0.03). We found the opposite association i.e., positive, between the ASD polygenic risk score and ERT (*b*=0.40; 95% CI: 0.10 to 0.70); however, this association varied across different p-value thresholds so should be interpreted with caution. We did not find evidence of a genetic correlation between ASD and ERT.

**Conclusion:** We found an observational association between poorer emotion recognition and increased autistic traits. Our genetic analyses revealed an association between ASD polygenic risk and the ERT outcome, which may suggest a shared genetic aetiology between these or a potential causal pathway. Our results may inform interventions targeting emotion recognition.

## Introduction

Autism Spectrum Disorder (ASD) is a neurodevelopmental condition that manifests in childhood and is characterised by persistent difficulties in social communication and interaction and restricted and repetitive behaviours (Campisi, Imran, Nazeer, Skokauskas, & Azeem, 2018). Emotion recognition is thought to be impaired in individuals with ASD (Lozier, Vanmeter, & Marsh, 2014; Uljarevic & Hamilton, 2013), which may contribute to impairments in social function. However, the direction of effect is unclear; social autistic traits may contribute to deficits in emotion recognition, or these deficits may contribute to, or exacerbate social autistic traits. Since emotion recognition is a potentially modifiable target (Penton-Voak et al., 2019; Penton-Voak, Bate, Lewis, & Munafò, 2012; Rawdon et al., 2018), a better understanding of its relationship with ASD and autistic traits, including the direction of association, may help inform management and treatment of ASD.

There is inconsistency in the literature on the association between ASD or autistic traits and emotion recognition of facial expressions. Numerous studies have found that those with ASD (or greater symptom severity) are less accurate and slower at identifying emotions than controls (or those with lower symptom severity) (Fridenson-Hayo et al., 2016; Griffiths et al., 2019; Loth et al., 2018; Lozier et al., 2014; Rump, Giovannelli, Minshew, & Strauss, 2009; Uljarevic & Hamilton, 2013; Wallace et al., 2011; Wingenbach, Ashwin, & Brosnan, 2017; Xavier et al., 2015). However, others have failed to show evidence of an association (Evers, Kerkhof, Steyaert, Noens, & Wagemans, 2014; Fink, De Rosnay, Wierda, Koot, & Begeer, 2014; Russo-Ponsaran et al., 2019; Tracy, Robins, Schriber, & Solomon, 2011). Although these studies have sought to establish whether an association between ASD and emotion recognition deficits exist, they have all been hampered by the use of relatively small sample sizes, cross-sectional study designs, and lack of consideration of the direction of the association. In addition, while these studies focused on individuals with a diagnosis of ASD it is also valuable to examine this relationship in non-clinical populations, as this presents the opportunity to assess how emotion recognition ability varies across a broader continuum of autistic traits.

We examined the association, and bidirectionality of the association, between autistic traits and emotion recognition using data from a large prospective birth cohort, the Avon Longitudinal Study of Parents and Children (ALSPAC). We conducted the study in three parts. First, we performed observational analyses to investigate the association between a continuous measure of autistic traits at age 8 and age 10 and performance on two distinct emotion recognition tasks at age 24 (Study 1) and age 8 (Study 2), respectively. Next, in an effort to strengthen the evidence, we performed polygenic score association analyses assessing the likely direction of this association further (Study 3). The aim of this study was to identify the direction between autistic traits and emotion recognition.

## Methods

### Cohort description

We included children from the ALSPAC birth cohort in this study. ALSPAC initially recruited 14,541 pregnant women residing in Avon, UK, with expected dates of delivery between 1st April 1991 and 31st December 1992. Of these initial pregnancies, 13,988 children were alive at age 1. When children were approximately age 7, additional eligible cases who had failed to join the study originally were recruited. As a result, there are data available for more than the 14,541 pregnancies mentioned above after age 7. There is a total sample size, for analyses using any data after age 7, of 14,901. Further details of the cohort and enrolment can be found in the cohort profile papers (Boyd et al., 2013; Fraser et al., 2013; Northstone et al., 2019). Please note that the study website contains details of all the data that are available through a fully searchable data dictionary and variable search tool (http://www.bristol.ac.uk/alspac/researchers/our-data/). Data collection from questionnaires or clinics from 2014 onwards was collected and managed by REDcap electronic data capture tools hosted at the University of Bristol (Harris et al., 2009, 2019). Ethics approval for the study was obtained from the ALSPAC Ethics and Law Committee and the Local Research Ethics Committees. Informed consent for the use of data collected via questionnaires and clinics was obtained from participants following the recommendations of the ALSPAC Ethics and Law Committee at the time.

### Phenotypic measures

#### Autistic traits

We used parent reported questionnaire responses from the Social and Communication Disorders Checklist (SCDC) (Skuse, Mandy, & Scourfield, 2005) total scores at two time points with mean ages of 7.66 (SD=0.14) and 10.72 (SD=0.14). These time points were selected to allow the association in both directions to be examined. The age 8 measure is the earliest time point at which the SCDC is available and therefore we have used this to avoid drop-out at later time points. However, the age 10 measure is more appropriate to assess the association with the emotion recognition task measured at age 8 for the direction of earlier emotion recognition ability and later autistic traits. The SCDC is a 12-item screening questionnaire for social and communication difficulties. Although it does not directly assess ASD, it is a good indicator of symptoms. Response options were ‘Not true’, ‘Quite/Sometimes true’ or ‘Very/Often true’, ranging from zero to two, with a maximum score of 24. Data was available for 7,807 participants at age 8 (51% were male) with a mean score of 2.80 (SD=3.73) and for 7,463 participants at age 10 (51% were male) with a mean score of 2.37 (SD=3.61). The distribution of scores is presented in Supplementary FigS1. As there is a zero-skewed distribution for both time points we also report the median score which is 2.00 (interquartile range [IQR]=4.00) at age 8 and 1.00 (IQR=3.00) at age 10. We have opted to use this continuous measure of social aspects of autistic traits (Bölte, Westerwald, Holtmann, Freitag, & Poustka, 2011) so that results are more generalisable to the wider population, but note that the prevalence of ASD diagnosis in the ALSPAC cohort is approximately 51.1 per 10,000 at age 11 (Williams, Thomas, Sidebotham, & Emond, 2008).

We also used data for sensitivity analyses from a questionnaire that mothers completed when the child was around age 9 which included a question asking whether the mother had been told that their child has autism or Asperger’s.

#### Emotion recognition at age 8

We used data from clinic visits (mean age=8.65 [SD=0.32]) for the facial expression subtest of the Diagnostic Analysis of Non-Verbal Accuracy (DANVA) (Nowicki & Duke, 1994). A series of 24 images of child faces expressing different emotions were shown and the child was asked to select which emotion they would assign to this face (i.e., happy, sad, angry or fearful). Facial expressions were presented in low or high intensity. Images were shown on a computer screen for two seconds, or if this was not available a manual version was used. Participants indicated their response and the tester clicked this option on the screen or noted this down. The number of errors for each emotion were also recorded. In this study we calculated the number of correct responses for each emotion, by subtracting the number of errors and missing data from the total number of tests. Data was available for 6,714 participants (50% were male) with a mean score of 19.39 (SD=2.78). The distribution of the number of correct responses for the DANVA is presented in Supplementary FigS2.

#### Emotion recognition at age 24

Data from clinic visits (mean age=24.46 [SD=0.78]) for the Emotion Recognition Task (ERT) (Attwood et al., 2017; Penton-Voak et al., 2012), from the ‘E-Prime’ session comprising three cognitive tasks, were used. During the ERT, participants were asked to assign emotions to facial images (i.e., happy, sad, angry, disgusted, surprised and fearful). Faces were shown on a screen for 0.2 seconds and then the participant selected one of the six emotions. There were eight levels of intensity for each emotion and each was presented twice, resulting in a total of 96 trials. We used the number of correct responses in analyses. Data was available for 3,562 participants (37% were male) with a mean score of 66.38 (SD=7.89). The distribution of the number of correct responses in the ERT is shown in Supplementary FigS3.

#### N-back working memory task

To assess whether any association between autistic traits and emotion recognition might be due to deficits in working memory, as opposed to deficits specific to emotion recognition, we also tested the association between autistic traits and working memory, as a sensitivity analysis. A working memory measure was also obtained for participants at the age 24 clinic: the *N*-back task (2-back condition) (Kirchner, 1958). During this task participants were presented with a series of numbers from zero to nine on a computer screen. They were asked to press the ‘1’ key when a number presented was the same as the number presented two trials previously, or a ‘2’ key if this was not the case. Overall performance on this task was determined with a measure of discriminability (*d’*), where a higher score indicates greater accuracy on the task. Further detail on this measure can be found in the Supplementary Material.

#### Stop signal task

Similarly, to assess whether any association might be due to reaction time, as opposed to deficits specific to emotion recognition, we also tested the association between autistic traits and reaction time in a stop signal task, as a sensitivity analysis. At the same clinic, participants were asked to complete the stop signal task (Logan, Cowan, & Davis, 1984). Participants were presented with the letter’s ‘X’ or ‘O’ sequentially on a computer screen and they were asked to press the relevant keys when the corresponding letter appeared, unless the stop signal tone was heard at which point, they should stop responding. There was a total of 256 trials, with four trials without the stop signal for every trial with the signal. Mean response times were calculated for the stop signal reaction time (SSRT) and used in our analyses as an indicator of response inhibition. Further detail on this measure can be found in the Supplementary Material.

#### Potential confounders

We included a number of potential confounders in the observational analyses between autistic traits and emotion recognition at age 24 (Study 1) and between emotion recognition at age 8 and later autistic traits (Study 2). These are shown on a timeline in Figure 1. We selected these potential confounders based on established risk factors for ASD and those that may theoretically be predictive of emotion recognition ability, based on the existing literature. First, we examined the unadjusted association between ASD and the outcome or exposure of interest (Model 1). Second, we included child’s sex, measures of socioeconomic status which included highest education and highest social class of mother and partner, income and tenure (Kelly et al., 2019; Rai et al., 2012) (Model 2). Third, we further included a measure for the child from the Wechsler Intelligence Scale for Children (WISC) (Oliveras-Rentas, Kenworthy, Roberson, Martin, & Wallace, 2012) and experience of head injuries assessed prior to the exposure (Milders, 2018) (Model 3). Fourth, we further included binary measures for any depression and any anxiety disorders (Attwood et al., 2017; Penton-Voak, Munafò, & Looi, 2017; Strang et al., 2012) from the Development and Well-Being Assessment (DAWBA) (Goodman, Ford, Richards, Gatward, & Meltzer, 2000) (Model 4). Further detail on these can be found in the Supplementary Material.

**Figure 1.**
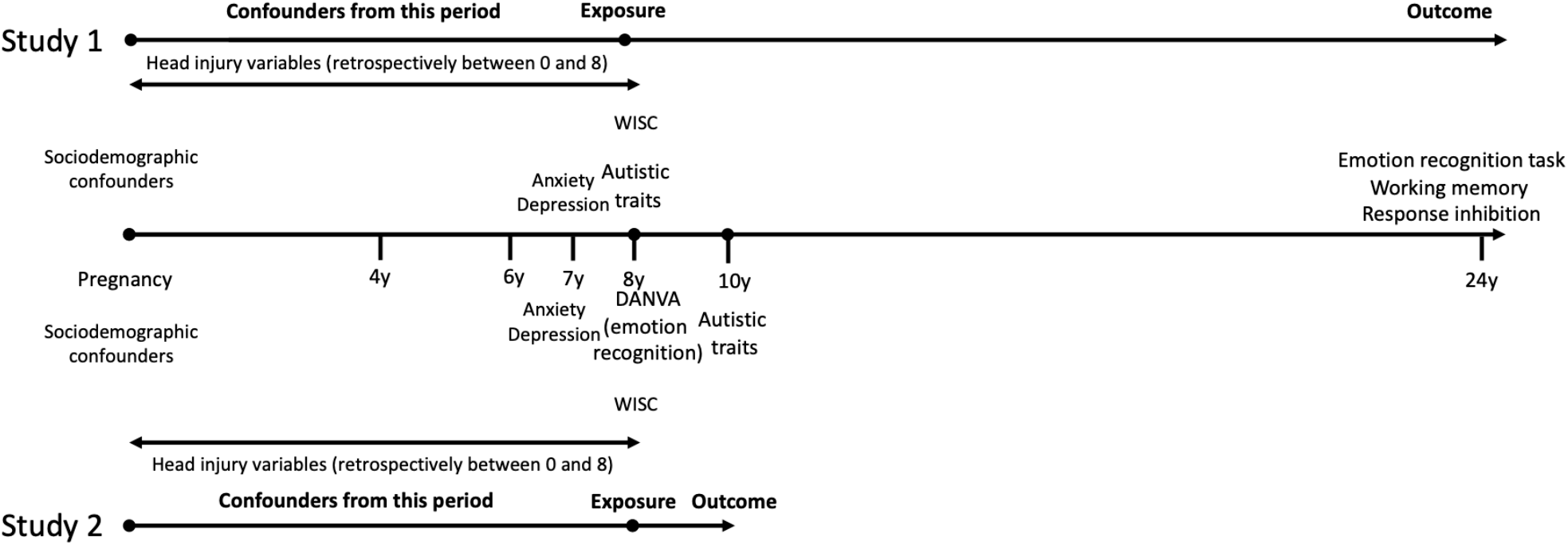
Timeline of exposure, outcome and potential confounders for Studies 1 and 2.

### Missing data

To ensure analyses provided sufficient power to detect associations, we imputed missing data for the exposure and covariates up to the number that data were available for the outcome. Supplementary FigS4 and S5 show the sample attrition relevant to Studies 1 and 2, respectively. Data for the SSRT and working memory cognitive measures were collected at the same time as the ERT measure and we imputed to the number where data was available for the ERT measure (N=3,562). Where autistic traits was the outcome we imputed up to the number who had data available for at least one ASD measure at ages 8 and 10 (N=9,071). Data were imputed in Stata version 15 using the *ice* and *match* commands. Further detail on the variables used for imputation is presented in the Supplementary Material.

### Genetic data

Children’s genetic data were obtained from blood samples collected during clinic visits. Samples were genotyped using the Illumina HumanHap 550 quad chip. Genome-wide data were generated by Sample Logistics and Genotyping Facilities at the Wellcome Trust Sanger Institute and LabCorp (Laboratory Corporation of America) using support from 23andMe. Quality control measures were used, and individuals excluded based on gender mismatches, minimal or excessive heterozygosity, disproportionate missingness, insufficient sample replication and being of non-European ancestry. Data from 9,115 children and 500,527 SNPs passed filters and data was imputed with a phased version of the 1000genomes reference panel from the Impute2 reference data repository. After these procedures, removing participants with cryptic relatedness >5% and those who had withdrawn consent, there were 8,252 children with genotype data available.

### Polygenic risk score

We constructed thirteen different polygenic risk scores using Plink (version 2) (Purcell et al., 2007) for each individual in ALSPAC using summary statistics from GWAS for ASD (Grove et al., 2019). We used genetic data in ALSPAC filtered by an imputation score of 0.8 and a minor allele frequency of 0.01. After filtering and selecting SNPs where genotype data was available, SNPs were clumped for linkage disequilibrium in Plink, using an R^2^ of 0.1. We generated weighted polygenic risk scores for ASD by summing the number of risk alleles present for each SNP (0, 1 or 2) weighted by the beta of that SNP from the discovery sample, using the --score command in Plink. Further details can be found in the Supplementary Material. Polygenic risk scores were z-standardised; therefore, results can be interpreted as per standard deviation (SD) increase in score.

### Statistical analysis

#### Observational associations

Analyses were conducted in R version 3.5.1 using linear regression models and quasi-Poisson models, where autistic traits were the outcome. The reported effect estimates are unstandardised coefficients. The libraries “haven”, “mice”, “dplyr” and “tidyr” were required to read in and use the imputed data from Stata. First, the associations between autistic traits at age 8 years and ERT, response inhibition and working memory outcomes at age 24 years were examined (Study 1). Next, we tested the association between emotion recognition at age 8 years and autistic traits at age 10 years (Study 2). For each of these associations we ran four different models adjusting for different numbers of potential confounders, as described above. Models using imputed data are reported in the main text. Non-imputed models are reported in the Supplementary Material.

#### Sensitivity analyses

We tested whether any associations between autistic traits and emotion recognition were due to more general cognitive function, using linear regression models including working memory and response inhibition as outcomes. If we found an association with either of these, we ran an additional model for Study 1 to include these as additional covariates, in order to see whether the emotion recognition association is independent of a more general cognitive deficit. Fully adjusted models for Study 1 and Study 2 excluding children with a diagnosis of autism at age 9 were also conducted.

#### Association between polygenic risk scores and phenotypes

For Study 3, we investigated the association between the ASD polygenic risk scores with outcomes for the ERT and DANVA. We also include results for associations with the SCDC autistic traits measure at age 8. We used linear regression models for the ERT and DANVA outcomes and a quasi-Poisson regression model for the autistic traits outcome. All models adjusted for child’s sex and the first 10 principal components (PCs).

#### Genetic correlation

In Study 3, we also used summary statistics from GWAS for ASD, the DANVA proportion index and the number of correct responses in the ERT to estimate genetic correlations between these phenotypes using linkage disequilibrium score (LDSC) regression. However, we were unable to calculate genetic correlations with the DANVA proportion index as the SNP heritability was too small.

## Results

### *Observational associations between autistic* traits *and emotion recognition at age 24 using imputed data (Study 1)*

Results are shown for autistic traits only, for each level of adjustment for Study 1 (Table 1). There was a negative association between autistic traits and the total number of correct emotion recognition responses in the fully adjusted model (*b*=-0.18; 95% CI: −0.27 to −0.09; *p*=8.31×10^−05^). These results are similar to the non-adjusted imputed results, although the effect is slightly attenuated. Results from the complete case non-imputed model are consistent (Supplementary Table S1). Supplementary Fig S6 demonstrates comparable distributions of the imputed and original variables. The results for Study 1 did not change when participants with a diagnosis of autism were excluded (Supplementary Table S2).

**Table 1.** Associations between autistic traits at age 8 and emotion recognition at age 24 (N=3,562)

| Model | Beta | 2.5% CI | 97.5% CI | P |
| --- | --- | --- | --- | --- |
| **Model 1<sup>a</sup>** | -0.27 | -0.36 | -0.18 | 5.21x10<sup>-09</sup> |
| **Model 2<sup>b</sup>** | -0.18 | -0.27 | -0.09 | 8.31x10<sup>-05</sup> |
| **Model 3<sup>c</sup>** | -0.17 | -0.26 | -0.08 | 1.90x10<sup>-04</sup> |
| **Model 4<sup>d</sup>** | -0.18 | -0.27 | -0.09 | 8.31x10<sup>-05</sup> |

*Models with number of correct responses in the emotion recognition task as the outcome and autistic traits as the exposure were adjusted as follows, in addition to covariates in the previous model. a: No covariates, b: sex + highest education + highest social class + income + tenure, c: WISC + head injury, d: depression + anxiety*

### Sensitivity analyses for Study 1

Results for the models with response inhibition and working memory as outcomes are shown in Supplementary Tables S3 and S4. There was evidence of a negative association between autistic traits and working memory in the fully adjusted model (*b*=-0.01; 95% CI: −0.02 to −0.003; *p*=0.01). However, there was no evidence of an association between autistic traits and response inhibition. We therefore further adjusted for working memory. The association of autistic traits at age 8 with the later ERT outcome remained even after adjustment (*b*=-0.16; 95% CI: −0.24 to −0.07; *p*=4.86×10^−04^).

### *Observational associations between* emotion recognition at age 8 *and* autistic traits *using imputed data (Study 2)*

Results from Study 2 are shown in Table 2. Results are shown for emotion recognition at age 8 only for each level of adjustment. There was a negative association between the total number of correct responses at age 8 and autistic traits at age 10 in the fully adjusted model (*b*=-0.04; 95% CI: −0.05 to 0.03; *p*=1.68×10^−10^). Results from the complete case non-imputed model are consistent (Supplementary Table S5). The results for Study 2 did not change when participants with a diagnosis of autism were excluded (Supplementary Table S6).

**Table 2.** Associations between emotion recognition at age 8 and autistic traits at age 10 (N=9,071)

| Model | Beta | 2.5% CI | 97.5% CI | P |
| --- | --- | --- | --- | --- |
| **Model 1<sup>a</sup>** | -0.06 | -0.07 | -0.05 | 6.11x10<sup>-20</sup> |
| **Model 2<sup>b</sup>** | -0.05 | -0.07 | -0.04 | 1.90x10<sup>-28</sup> |
| **Model 3<sup>c</sup>** | -0.04 | -0.05 | -0.03 | 2.20x10<sup>-10</sup> |
| **Model 4<sup>d</sup>** | -0.04 | -0.05 | -0.03 | 1.68x10<sup>-10</sup> |

*Models with the autistic traits as the outcome and number of correct responses in the DANVA as the exposure were adjusted as follows, in addition to covariates in the previous model. a: No covariates, b: sex + highest education + highest social class + income + tenure, c: WISC + head injury, d: depression + anxiety*

### Association between polygenic risk scores and phenotypes (Study 3)

Results from linear regression models between polygenic risk scores for ASD and outcomes for autistic traits, the ERT and DANVA are shown in Supplementary Tables S7, S8 and S9 and visualised in supplementary Figures S8, S9 and S10, respectively. The ASD polygenic risk score is associated with the SCDC measure at age 8 for the polygenic risk scores constructed with a p-value greater than and including 0.1 (*b*=0.05; 95% CI: 0.01 to 0.08; p=0.01). The ASD polygenic risk score was not consistently associated with the ERT outcome and where there was evidence of an association this was in the opposite direction to what we observe phenotypically, where an increased genetic risk for ASD is associated with a greater number of correct responses in the ERT. The strongest association we found was with a p-value threshold of 0.001 (*b*=0.40; 95% CI: 0.10 to 0.70, p=0.01). However, for those results without evidence of an association the direction of effect is opposite. We do not observe any associations between the ASD polygenic risk score and the DANVA outcome. Finally, we did not find evidence of a genetic correlation between ERT responses and ASD (rG=0.30, SE=0.21, *p*=0.15).

## Discussion

We examined the association between autistic traits and emotion recognition in a large population-based birth cohort. Higher autistic traits at age 8 were associated with poorer emotion recognition at age 24. In addition, better emotion recognition at age 8 was associated with lower autistic traits at age 10. We also examined the association between response inhibition and working memory with autistic traits. We did not find any association with response inhibition; however, we did find an association with working memory. Nevertheless, even after adjusting for working memory, the association between autistic traits at age 8 and ERT performance at age 24 remained, suggesting these emotion recognition deficits are at least partially independent from any general cognitive deficit. We also found that excluding participants with a diagnosis of autism had minimal impact on the results, indicating these results are generalisable to non-clinical populations.

### Comparison with previous literature

Our findings are in line with previous studies which demonstrate a negative association between ASD (or greater symptom severity) and emotion recognition (Fridenson-Hayo et al., 2016; Griffiths et al., 2019; Loth et al., 2018; Lozier et al., 2014; Rump et al., 2009; Uljarevic & Hamilton, 2013; Wallace et al., 2011; Wingenbach et al., 2017; Xavier et al., 2015). A lack of evidence of this association in other studies (Evers et al., 2014; Fink et al., 2014; Russo-Ponsaran et al., 2019; Tracy et al., 2011) may be due to much smaller sample sizes and lower power in these studies, compared to ours, and/or reliance on cross-sectional designs. Utilising longitudinal data from a large prospective cohort, provides robust evidence of an association between poorer emotion recognition and autistic traits. We increased our sample size and thus power by using multiple imputation on our data and also reduced biases that might be present when using complete case analyses. The tasks used in different studies also vary so it may be that some tasks are better designed to detect these effects than others. Our observational findings suggest that there is an association between earlier autistic traits and later emotion recognition deficits, and vice versa; therefore, it is unclear whether emotion recognition is a symptom or cognitive biomarker of ASD, or on the causal pathway from ASD or whether emotion recognition deficits exacerbate autistic traits. To the best of our knowledge, this is the first study to use this approach of observational and genetic techniques to examine this association.

We found, somewhat unexpectedly, that an increased genetic risk for ASD was associated with a greater number of correct responses in the emotion recognition task at age 24, but not at age 8. However, the lack of a consistent direction of effect across the polygenic risk scores means these results should be interpreted with caution. In addition, we have not corrected these results for multiple testing, because our focus is on estimating effect sizes, rather than declaring results as statistically significant or not. However, we recognise that we have conducted a large number of analyses, and these results should also be interpreted with appropriate caution. Only one other study, to our knowledge, has examined the association between ASD polygenic risk score and emotion recognition outcomes, and they did not find an association with total correct responses after adjusting for multiple testing (Wendt, Carvalho, Gelernter, & Polimanti, 2019). A previous study examining the genetic correlation between ASD and scores on a test looking at cognitive empathy also did not find evidence of an association, although this tests mental state recognition more broadly as opposed to the specific emotions assessed in the tasks we have included (Warrier et al., 2018). Therefore, further examination of this association would help to identify whether this is robust, and if so, what is the direction of effect. The small sample size used for the emotion recognition GWAS may indicate a lack of power to accurately detect this effect in our genetic correlation analysis. It would be useful to investigate this further, potentially with Mendelian randomisation type or genomic structural equation modelling (Grotzinger et al., 2019) approaches, but the current GWAS for emotion recognition are underpowered to do this. We were also unable to examine the association between emotion recognition polygenic risk scores and autistic traits due to sample overlap with the GWAS for the DANVA (Coleman et al., 2017) and ERT (Mahedy et al., 2019) also being conducted in ALSPAC. To investigate whether our findings could be a result of shared genetic aetiology we looked at genetic correlations between these outcomes. However, we were unable to examine this fully in our study due to poor genetic signal for the DANVA. We did not find evidence of a genetic correlation between ASD and number of correct responses in the ERT, but this may be due to the small sample size in the GWAS of emotion recognition.

### Limitations

Whilst we observe an association between autistic traits and emotion recognition in this study, there are a few considerations to take into account when interpreting these results. Firstly, as with any study there will be a degree of measurement error in the variables we used. Secondly, although we used longitudinal data to assess whether there was evidence of a possible direction of effect, it is possible that any effects of emotion recognition deficits on autistic traits or vice versa may occur at an earlier developmental stage than we used in this study. It would be useful to examine the direction of association in additional samples that have measures available at earlier developmental time points. However, given that autistic traits are relatively stable over time (Robinson et al., 2011), it is likely that the findings in this study would be similar if earlier measures were available. Thirdly, the accuracy of the polygenic risk scores can also be influenced by the target sample size, which is relatively small for the outcomes compared to the discovery sample size for ASD. In addition, the ASD polygenic risk score was only predictive of our SCDC measure at age 8 at less stringent p-value thresholds, although it is important to highlight that the original GWAS was conducted based on diagnosis and not a continuous measure. Conducting GWAS of emotion recognition in larger samples and perhaps continuous measures of ASD may give us greater power to detect effects in future studies.

### Conclusion

Our results indicate that there is an association between emotion recognition deficits and increased autistic traits. This is supported by our genetic analyses which revealed an association between polygenic risk scores for ASD and emotion recognition. Our genetic analyses were underpowered to identify a causal direction of effect, but suggest that there may be a shared genetic aetiology which could explain some of our observational findings, and could in principle reflect a potential causal pathway from ASD to emotion recognition. Given that emotion recognition is a modifiable target, this is worth exploring further. Future research would benefit from using better powered GWAS; therefore, there is a need to collect these data in larger samples. Our results may be useful in supporting development of interventions that target emotion recognition deficits in ASD.

## Data Availability

The ALSPAC data management plan (http://www.bristol.ac.uk/alspac/researchers/data-access/documents/alspac-data-management-plan.pdf) describes in detail the policy regarding data sharing, which is through a system of managed open access. The steps below highlight how to apply for access to the data included in this paper and all other ALSPAC data. The datasets used in this analysis are linked to ALSPAC project number B3113; please quote this project number during your application.
1. Please read the ALSPAC access policy (PDF, 627 kB) which describes the process of accessing the data and samples in detail, and outlines the costs associated with doing so.
2. You may also find it useful to browse the fully searchable ALSPAC research proposals database, which lists all research projects that have been approved since April 2011.
3. Please submit your research proposal for consideration by the ALSPAC Executive Committee. You will receive a response within 10 working days to advise you whether your proposal has been approved.
If you have any questions about accessing data, please.

## Acknowledgements

We are extremely grateful to all the families who took part in this study, the midwives for their help in recruiting them, and the whole ALSPAC team, which includes interviewers, computer and laboratory technicians, clerical workers, research scientists, volunteers, managers, receptionists and nurses.

## Funding

This work was supported in part by the UK Medical Research Council Integrative Epidemiology Unit at the University of Bristol (Grant ref: MC_UU_00011/1, MC_UU_00011/7). The UK Medical Research Council and Wellcome (Grant ref: 217065/Z/19/Z) and the University of Bristol provide core support for ALSPAC. This publication is the work of the authors and ZER, LM, AJ, GDS, IPV, ASA and MRM will serve as guarantors for the contents of this paper. A comprehensive list of grants funding is available on the ALSPAC website (http://www.bristol.ac.uk/alspac/external/documents/grant-acknowledgements.pdf). GWAS data was generated by Sample Logistics and Genotyping Facilities at Wellcome Sanger Institute and LabCorp (Laboratory Corporation of America) using support from 23andMe. ZER is supported by the Elizabeth Blackwell Institute for Health Research, University of Bristol and the Wellcome Trust Institutional Strategic Support Fund (Grant ref:204813/Z/16/Z). LM is supported by the Elizabeth Blackwell Institute for Health Research, University of Bristol and the Wellcome Trust Institutional Strategic Support Fund (Grant ref: 204813/Z/16/Z). IPV and MRM are supported by the National Institute for Health Research Bristol Biomedical Research Centre.

IPV and MRM are co-directors of Jericoe Ltd. a company that licenses software for the assessment and modification of emotion recognition. All other authors have declared that they have no competing or potential conflicts of interest.

## Key points

- Emotion recognition deficits are thought to be present in individuals with ASD which may impair social function, but findings are mixed.
- In our study we found that there was a negative association between ASD symptoms and both later and earlier emotion recognition.
- We also found a positive association between a polygenic risk score for ASD and emotion recognition ability.
- This may indicate that genetic variants for ASD are also shared with emotion recognition, or that ASD causally influences emotion recognition.
- Our findings may have implications for intervention development targeting emotion recognition, if this is in fact causal.

## Notes

### Author Declarations

Ethics approval for the study was obtained from the ALSPAC Ethics and Law Committee and the Local Research Ethics Committees. Informed consent for the use of data collected via questionnaires and clinics was obtained from participants following the recommendations of the ALSPAC Ethics and Law Committee at the time.

### Summary of Updates

We updated this paper by rerunning analyses with autistic traits measured at different time points. Study 1 now examines the association between autistic traits at age 8 and responses on an emotion recognition task at age 24 and study 2 examines the association between responses on an emotion recognition task at age 8 and autistic traits at age 10. We have included additional polygenic risk score analyses in the supplementary materials as well as some additional sensitivity analyses.

