## Supplementary materials for "Examining the bidirectional association between emotion recognition and autistic traits using observational and genetic analyses"

*N-back working memory task*

This task has been described previously by Mahedy and colleagues (Mahedy et al., 2019). In this task, participants continuously monitored a series of numbers (0-9) presented on a computer screen and pressed the ‘1’ key if the number was the same as the number presented two trials previously (i.e., 2-back), or the ‘2’ key if it was not. Stimuli numbers were presented in black on a white background with a random spatial jitter of 180 pixels in the y-axis and 200 pixels in the x-axis. Each target was presented for 500 ms, followed by a 3,000 ms window in which to respond. There was an initial practice block which consisted of 12 trials containing two targets. This was followed by an experimental block, consisting of 48 trials, with 8 targets, where the target was the number that was identical to the one presented 2 trials previously. Three outcomes were examined for the N-back task: (i) number of hits, or the percentage of matching numbers correctly identified as matches, (ii) false alarms, or the percentage of non-matching numbers incorrectly identified as matches, and (iii) discriminability index, d′, a signal-detection metric that takes into account both hits and false alarms to derive an overall estimate of signal-detection ability (McNicol, 1972; Stanislaw and Todorov, 1999). d′ was calculated as follows:

d’ = invnorm(hits) – invnorm(false alarms)

*Stop signal task*

This task has been described previously by Mahedy and colleagues (Mahedy et al., 2019). In this task, participants were asked to sit in front of a computer screen with their two index fingers placed on two stimulus keys, one for ‘X’ and one for ‘O’. There were two types of trial, the ‘go’ trials and the stop signal trials. In the ‘go’ trials, participants were asked to fixate on a plus sign (+) in the centre of the computer screen. An ‘X’ or ‘O’ was presented on the screen and the participant had to press the corresponding key as quickly as possible. On 25% of the trials, a beep is heard (stop signal), randomly after the ‘X’ or ‘O’ appears. Participants were asked not to press the corresponding key when they heard the stop signal/beep and to wait for the next trial to begin. If the beep was not heard the participant was asked to press the corresponding key according to what was presented on screen. There were 32 practice trials and the main task consisted of 256 trials, comprising 4 blocks of 64 trials. Each block of 64 trials consisted of 4 sub-blocks of 16 trials. Each sub-block consisted of 12 trials without a stop-signal and 4 trials with a stop-signal/beep. Mean response times for this were calculated. Four metrics were obtained for the stop signal task: (i) an estimate of stop signal reaction time (SSRT) was calculated and used as a measure of inhibitory control (shorter SSRTs indicate slower inhibition); (ii) ‘go’ reaction time; (iii) ‘go’ accuracy; and (iv) ‘stop’ accuracy. In our study we used the SSRT, which was calculated as follows:

SSRT_med_ = Go Reaction Time_med_ – Stop Signal Delay_med_

The Stop Signal Delay_med_ (SSD) was calculated using a weighted least squares linear regression, to predict SSD based on the probability of responding given a stop-signal. This was then used to estimate the SSD where the probability of the participant failing to inhibit was 50%.

*Potential confounders*

We selected potential confounders based on established risk factors for autism spectrum disorder (ASD) and those that may theoretically be predictive of emotion recognition ability. These included child’s sex, measures of socioeconomic status (highest education and social class of mother and partner, income and tenure), a measure from the Wechsler Intelligence Scale for Children (WISC), head injuries and binary measures for depression and anxiety.

Highest social class was based on occupation of the mother and partner from questionnaire data at approximately 32 weeks gestation. Highest education level for mothers and partners was taken from questionnaire data, where participants were asked “What educational qualifications do you, your partner, your mother, and your father have?”. They were asked to select all options that applied to them and we used the highest education qualification for the participant. The options for this were Certificate of Secondary Education (CSE) / none, vocational qualifications, O level (qualifications usually taken at age 16), A level (qualifications usually taken at age 18) or degree. We used the highest from mothers and partners if both were provided. for income (quintiles) and tenure (owned/mortgaged, subsidised renting and private renting) were collected during pregnancy, around the time of enrolment.

We used total scores from the WISC (Wechsler et al., 1992), assessed at age 8 in research clinics, as a measure of IQ. A short form of the measure was used, comprising of five verbal and five performance subtests, results of which were combined to give a total score for IQ. The depression and anxiety measures were obtained from child-based questionnaires completed by the mother/caregiver when the child was around age 8, where questions based on DSM-IV criteria were asked and a binary measure of any depressive disorder and any anxiety disorder were used. A head injury variable was created based on questionnaire data up to age 8.

*Multiple Imputation*

To impute the data to use for the emotion recognition task (ERT), SSRT and working memory analyses, we included all relevant outcome, exposure and covariate variables, as well as the Diagnostic Analysis of Non-Verbal Accuracy (DANVA) measure, working memory at age 11, measures of autistic traits also using the SCDC at ages 8, 10 and 13, a total score from the strengths and difficulties questionnaire (SDQ) at age 6, the Wechsler Abbreviated Scale of Intelligence (WASI) at age 15, the sociability scale of the Emotionality, Activity, Sociability (EAS) temperament scale at age 3, measures for crowding and parity. To impute the data to use for the Diagnostic Analysis of Nonverbal Accuracy (DANVA) analyses, we used the same variables.

*Polygenic risk scores*

We constructed thirteen different polygenic risk scores (PRS) based on different thresholds of the ASD genome-wide association study (GWAS) p-values (0.5, 0.4, 0.3, 0.2, 0.1, 0.05, 0.01, 1x10^-3^, 1x10^-4^, 1x10^-5^, 1x10^-6^, 1x10^-7^, 5x10^-8^). We have presented all results for these polygenic risk scores in supplementary Tables S1, S2 and S3 and supplementary Figures S8, S9 and S10).

**Supplementary Table S1. Unimputed result for the association between autistic traits at age 8 and the emotion recognition task outcome for the fully adjusted model (N=2,202).**

| **Beta** | **2.5% CI** | **97.5% CI** | **P** |
| --- | --- | --- | --- |
| -0.15 | -0.25 | -0.05 | 4.75x10^-03^ |

*Model with number of correct responses in the emotion recognition task as the outcome and autistic traits as the exposure adjusted for sex, highest education, highest social class, income, tenure, WISC, head injury, depression and anxiety*

**Supplementary Table S2. Associations between autistic traits at age 8 and emotion recognition at age 24 excluding participants with a diagnosis of autism for the fully adjusted model (N=3,534)**

| **Beta** | **2.5% CI** | **97.5% CI** | **P** |
| --- | --- | --- | --- |
| -0.19 | -0.27 | -0.10 | 1.54x10^-243^ |

*Models with number of correct responses in the emotion recognition task as the outcome and autistic traits as the exposure were adjusted as follows: sex + highest education + highest social class + income + tenure + WISC + head injury + depression + anxiety*

**Supplementary Table S3. Imputed results for associations between autistic traits at age 8 and SSRT outcome (N=3,562).**

| **Model** | **Beta** | **2.5% CI** | **97.5% CI** | **P** |
| --- | --- | --- | --- | --- |
| Model 1^a^ | 0.67 | 0.005 | 1.33 | 0.05 |
| Model 2^b^ | 0.73 | 0.06 | 1.39 | 0.03 |
| Model 3^c^ | 0.41 | -0.25 | 1.06 | 0.23 |
| Model 4^d^ | 0.44 | -0.23 | 1.11 | 0.19 |

*Models with SSRT as the outcome and autistic traits as the exposure were adjusted as follows in addition to covariates in the previous model. a: No covariates, b: sex + highest education + highest social class + income + tenure, c: WISC + head injury, d: depression + anxiety*

**Supplementary Table S4. Imputed results for associations between autistic traits at age 8 and working memory outcome (N=3,562).**

| **Model** | **Beta** | **2.5% CI** | **97.5% CI** | **P** |
| --- | --- | --- | --- | --- |
| Model 1^a^ | -0.02 | -0.03 | -0.01 | 5.38x10^-05^ |
| Model 2^b^ | -0.02 | -0.03 | -0.01 | 5.05x10^-05^ |
| Model 3^c^ | -0.01 | -0.02 | -0.004 | 5.65x10^-03^ |
| Model 4^d^ | -0.01 | -0.02 | -0.003 | 0.01 |

*Models with working memory as the outcome and autistic traits as the exposure were adjusted as follows in addition to covariates in the previous model. a: No covariates, b: sex + highest education + highest social class + income + tenure, c: WISC + head injury, d: depression + anxiety*

**Supplementary Table S5. Unimputed result for the associations between the DANVA measure and autistic traits at age 10 for the fully adjusted model (N=4,103).**

| **Beta** | **2.5% CI** | **97.5% CI** | **P** |
| --- | --- | --- | --- |
| -0.04 | -0.05 | -0.02 | 3.90x10^-06^ |

*Models with the autistic traits as the outcome and number of correct responses in the DANVA as the exposure adjusted for sex, highest education, highest social class, income, tenure, WISC, head injury, depression and anxiety*

**Supplementary Table S6. Associations between emotion recognition at age 8 and autistic traits at age 10 excluding participants with a diagnosis of autism for the fully adjusted model (N=8,984)**

| **Beta** | **2.5% CI** | **97.5% CI** | **P** |
| --- | --- | --- | --- |
| -0.04 | -0.05 | -0.02 | 3.03x10^-08^ |

*Models with the autistic traits as the outcome and number of correct responses in the DANVA as the exposure were adjusted as follows: sex + highest education + highest social class + income + tenure + WISC + head injury + depression + anxiety*

**Supplementary Table S7. Associations between standardised ASD polygenic risk score and autistic traits at age 8 in ALSPAC (N=5,306).**

| **P-value threshold for polygenic risk score** | **Beta** | **2.5% CI** | **97.5% CI** | **P** | **R^2^** |
| --- | --- | --- | --- | --- | --- |
| 0.00000005 | 0.02 | -0.01 | 0.06 | 0.18 | 4.45X10^-04^ |
| 0.0000001 | 0.02 | -0.01 | 0.06 | 0.18 | 4.45X10^-04^ |
| 0.000001 | -0.02 | -0.05 | 0.02 | 0.39 | 5.48X10^-05^ |
| 0.00001 | 0.01 | -0.03 | 0.04 | 0.77 | 2.74X10^-05^ |
| 0.0001 | 0.01 | -0.02 | 0.05 | 0.56 | 8.40X10^-05^ |
| 0.001 | 0.01 | -0.02 | 0.05 | 0.46 | 1.51X10^-04^ |
| 0.01 | 0.02 | -0.01 | 0.06 | 0.16 | 2.98X10^-04^ |
| 0.05 | 0.03 | -0.003 | 0.07 | 0.07 | 5.74X10^-04^ |
| 0.1 | 0.05 | 0.01 | 0.08 | 0.01 | 1.23X10^-03^ |
| 0.2 | 0.05 | 0.01 | 0.08 | 0.01 | 1.26X10^-03^ |
| 0.3 | 0.05 | 0.01 | 0.08 | 0.01 | 1.15X10^-03^ |
| 0.4 | 0.04 | 0.01 | 0.08 | 0.01 | 1.05X10^-03^ |
| 0.5 | 0.05 | 0.01 | 0.08 | 0.01 | 1.14X10^-03^ |

*Models with the autistic traits as the outcome and the 13 different autism spectrum disorder (ASD) polygenic risk score as the exposure adjusted for sex and the first 10 principal components.*

**Supplementary Table S8. Associations between standardised ASD polygenic risk score and the ERT outcome at age 24 in ALSPAC (N=2,555).**

| **P-value threshold for polygenic risk score** | **Beta** | **2.5% CI** | **97.5% CI** | **P** | **R^2^** |
| --- | --- | --- | --- | --- | --- |
| 0.00000005 | -0.12 | -0.42 | 0.18 | 0.45 | 3.50X10^-04^ |
| 0.0000001 | -0.12 | -0.42 | 0.18 | 0.45 | 3.50X10^-04^ |
| 0.000001 | -0.07 | -0.37 | 0.23 | 0.64 | 1.34X10^-04^ |
| 0.00001 | -0.16 | -0.47 | 0.14 | 0.29 | 4.99X10^-04^ |
| 0.0001 | -0.01 | -0.32 | 0.30 | 0.95 | 6.33X10^-06^ |
| 0.001 | 0.40 | 0.10 | 0.70 | 0.01 | 2.45X10^-03^ |
| 0.01 | 0.35 | 0.05 | 0.65 | 0.02 | 1.99X10^-03^ |
| 0.05 | 0.12 | -0.18 | 0.42 | 0.43 | 2.48X10^-04^ |
| 0.1 | 0.09 | -0.22 | 0.39 | 0.56 | 1.17X10^-04^ |
| 0.2 | 0.02 | -0.28 | 0.33 | 0.87 | 8.27X10^-06^ |
| 0.3 | -0.004 | -0.31 | 0.30 | 0.98 | 4.00X10^-07^ |
| 0.4 | -0.002 | -0.31 | 0.30 | 0.99 | 1.12X10^-07^ |
| 0.5 | 0.002 | -0.30 | 0.30 | 0.99 | 1.09X10^-07^ |

*Models with the emotion recognition task (ERT) measure as the outcome and the 13 different autism spectrum disorder (ASD) polygenic risk score as the exposure adjusted for sex and the first 10 principal components.*

**Supplementary Table S9. Associations between standardised ASD polygenic risk score and the DANVA at age 8 in ALSPAC (N=4,901).**

| **P-value threshold for polygenic risk score** | **Beta** | **2.5% CI** | **97.5% CI** | **P** | **R^2^** |
| --- | --- | --- | --- | --- | --- |
| 0.00000005 | 0.002 | -0.07 | 0.08 | 0.96 | 7.14X10^-08^ |
| 0.0000001 | 0.002 | -0.07 | 0.08 | 0.96 | 7.14X10^-08^ |
| 0.000001 | -0.01 | -0.09 | 0.07 | 0.80 | 2.63X10^-05^ |
| 0.00001 | -0.01 | -0.09 | 0.06 | 0.76 | 2.54X10^-05^ |
| 0.0001 | -0.01 | -0.09 | 0.07 | 0.82 | 1.57X10^-05^ |
| 0.001 | -0.001 | -0.08 | 0.07 | 0.99 | 2.93X10^-06^ |
| 0.01 | 0.01 | -0.06 | 0.09 | 0.73 | 1.53X10^-05^ |
| 0.05 | -0.02 | -0.10 | 0.06 | 0.59 | 7.09X10^-05^ |
| 0.1 | -0.01 | -0.09 | 0.06 | 0.75 | 3.49X10^-05^ |
| 0.2 | 0.0005 | -0.08 | 0.08 | 0.99 | 9.59X10^-07^ |
| 0.3 | 0.002 | -0.07 | 0.08 | 0.96 | 2.59X10^-08^ |
| 0.4 | 0.003 | -0.07 | 0.08 | 0.94 | 2.66x10^-11^ |
| 0.5 | -0.003 | -0.08 | 0.07 | 0.94 | 4.88X10^-06^ |

*Models with the Diagnostic Analysis of Nonverbal Accuracy (DANVA) measure as the outcome and the 13 different autism spectrum disorder (ASD) polygenic risk score as the exposure adjusted for sex and the first 10 principal components.*

**Supplementary Figure S1. Histogram of SCDC assessed autistic traits at ages 8 and 10***The distribution of SCDC assessed autistic traits at ages 8 and 10 indicates that these data are zero-skewed.*

SCDC score at age 10

SCDC score at age 8

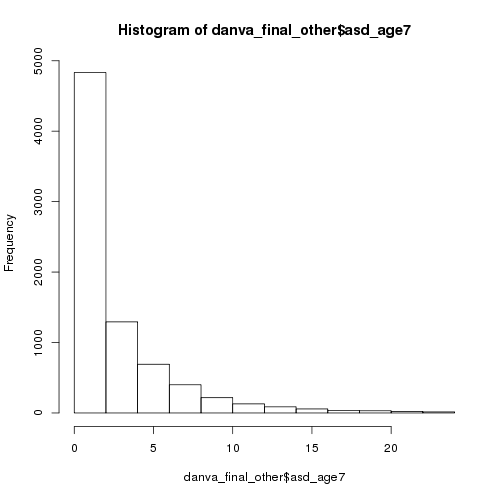

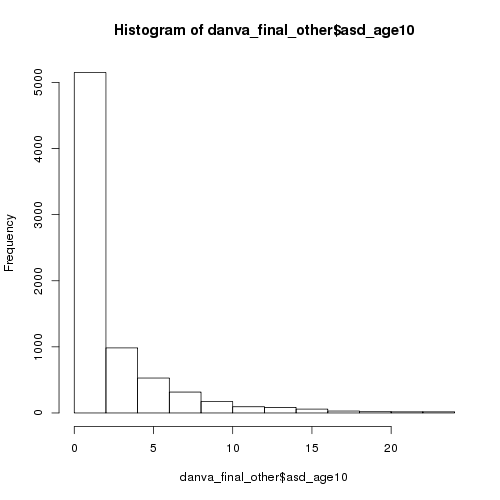

**Supplementary Figure S2. Histogram of number of correct responses in the DANVA.**

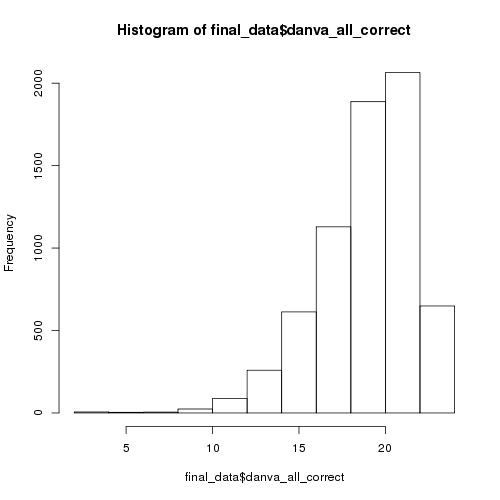

Number of correct responses in the DANVA

*The distribution of the number of correct responses in the DANVA indicates that these data are slightly negatively skewed.*

**Supplementary Figure S3. Histogram of number of correct responses in the ERT.**

Number of correct responses in the ERT

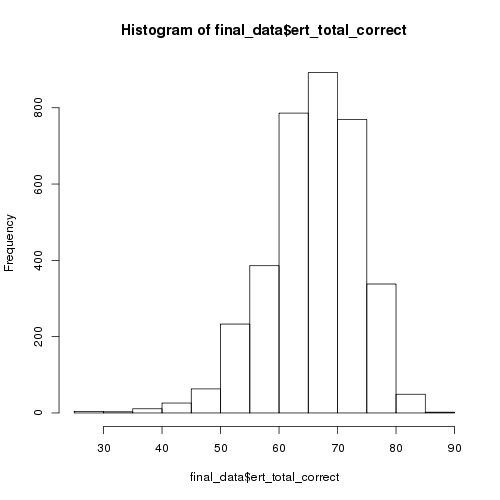

*The distribution of the number of correct responses in the DANVA indicates that these data are slightly negatively skewed.*

**Supplementary Figure S4. Sample attrition in ALSPAC for measures at the age 24 clinic (Study 1).**

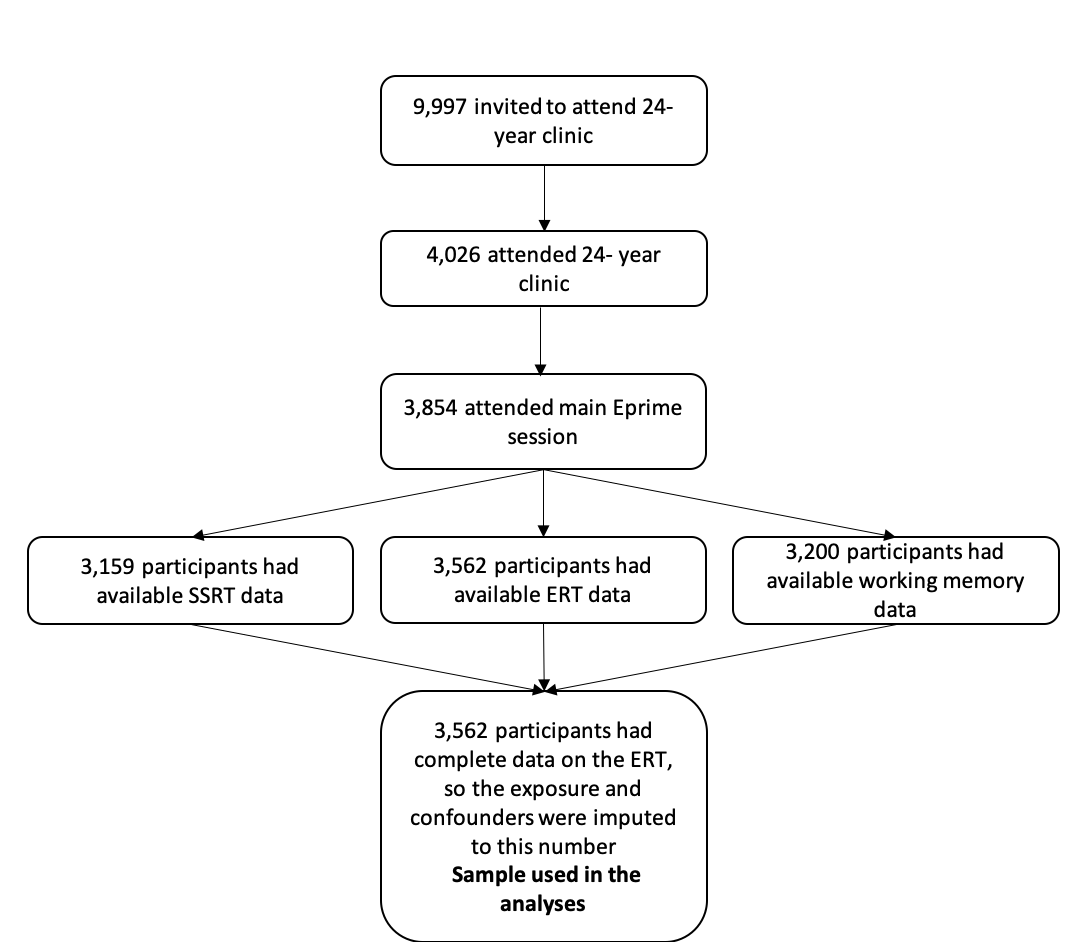

**Supplementary Figure S5. Sample attrition in ALSPAC for the autistic traits outcome at age 10 (Study 2).**

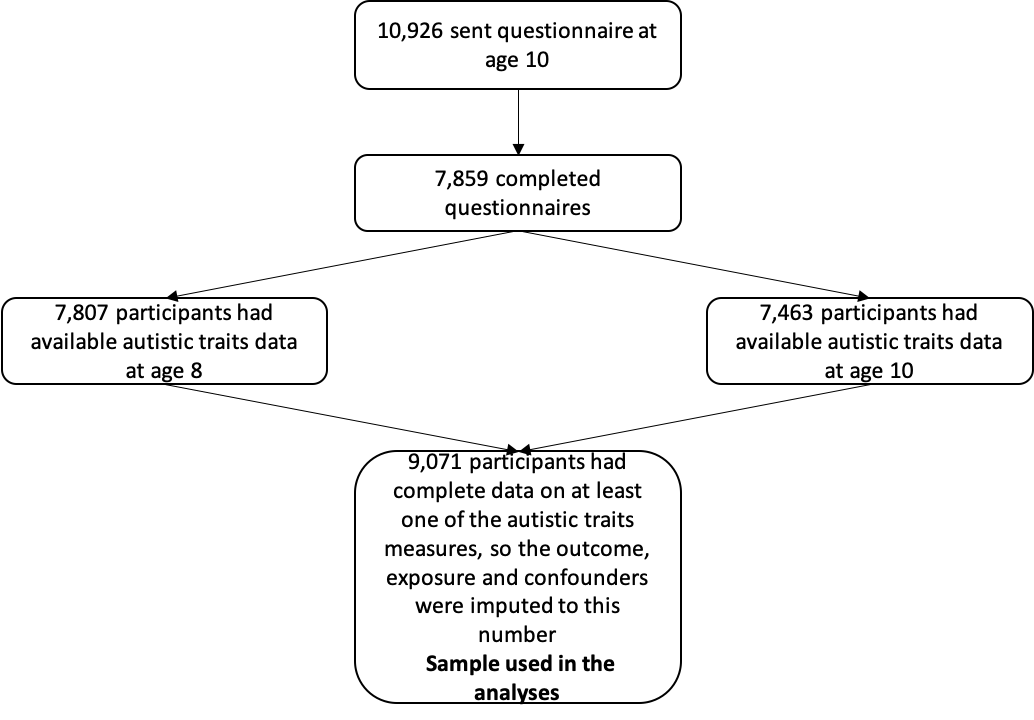

**Supplementary Figure S6. Distribution of imputed variables compared to original data for ERT, SSRT and working memory outcome analyses.**

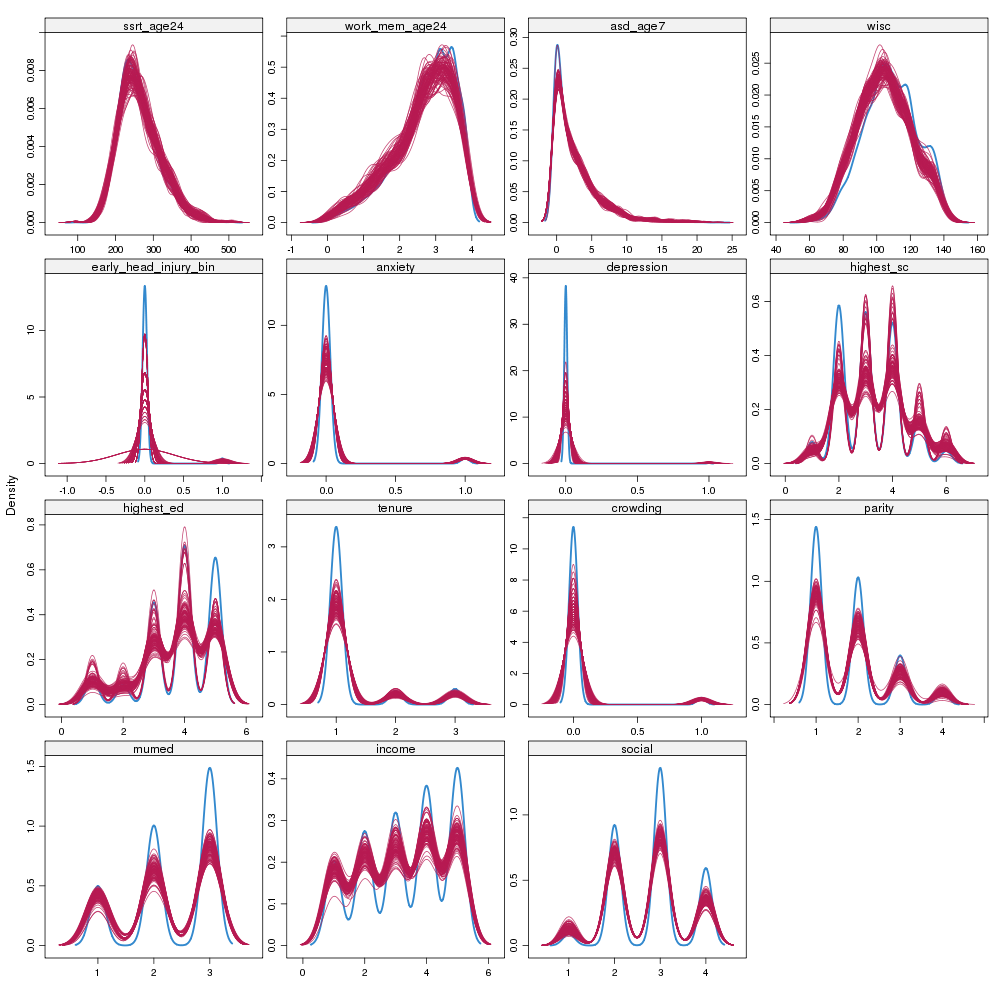
*Density plots for each variable that was imputed for the ERT, SSRT, working memory and autistic traits at age 8 analyses. Blue represents the observed data and red represents the imputed data for 100 imputations.*

**Supplementary Figure S7. Distribution of imputed variables compared to original data for autistic traits at age 10 outcome analyses.**

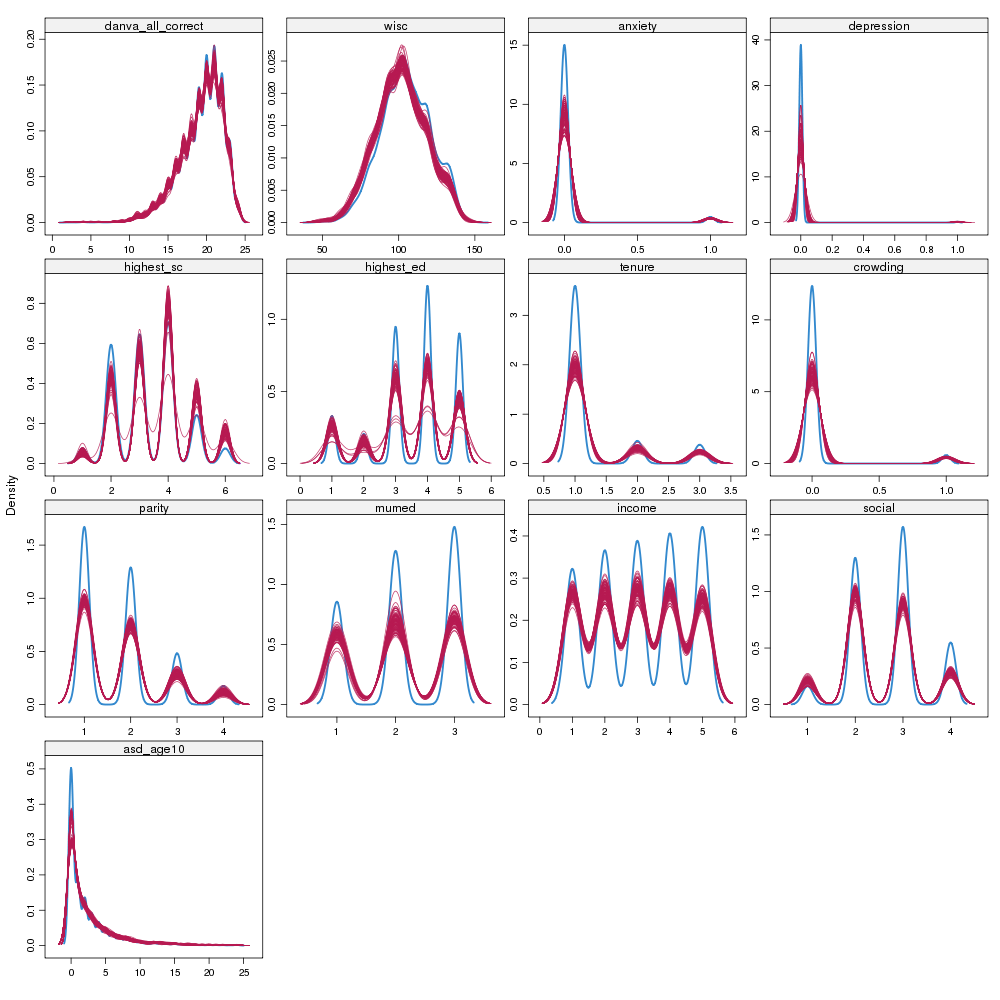
*Density plots for each variable that was imputed for the DANVA and autistic traits at age 10 analyses. Blue represents the observed data and red represents the imputed data for 100 imputations.*

**Supplementary Figure S8. Plot of associations between standardised ASD polygenic risk score and autistic traits at age 8 in ALSPAC.**

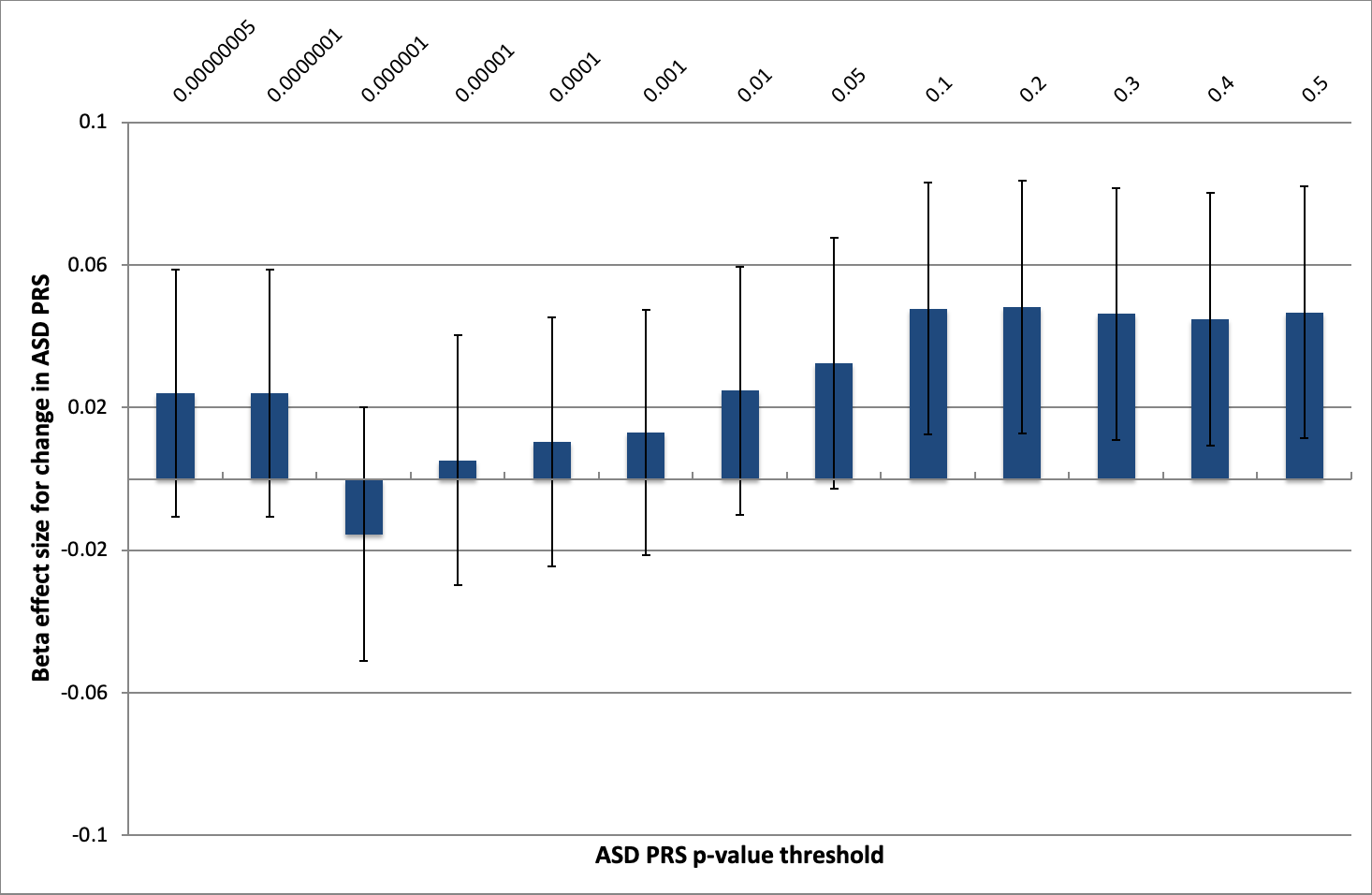

*The bars represent the effect size of the association with 95% confidence intervals shown. PRS=polygenic risk score, ASD=autism spectrum disorder, ALSPAC=Avon longitudinal study of parents and children*

**Supplementary Figure S9. Plot of associations between standardised ASD polygenic risk score and the ERT outcome at age 24 in ALSPAC.**

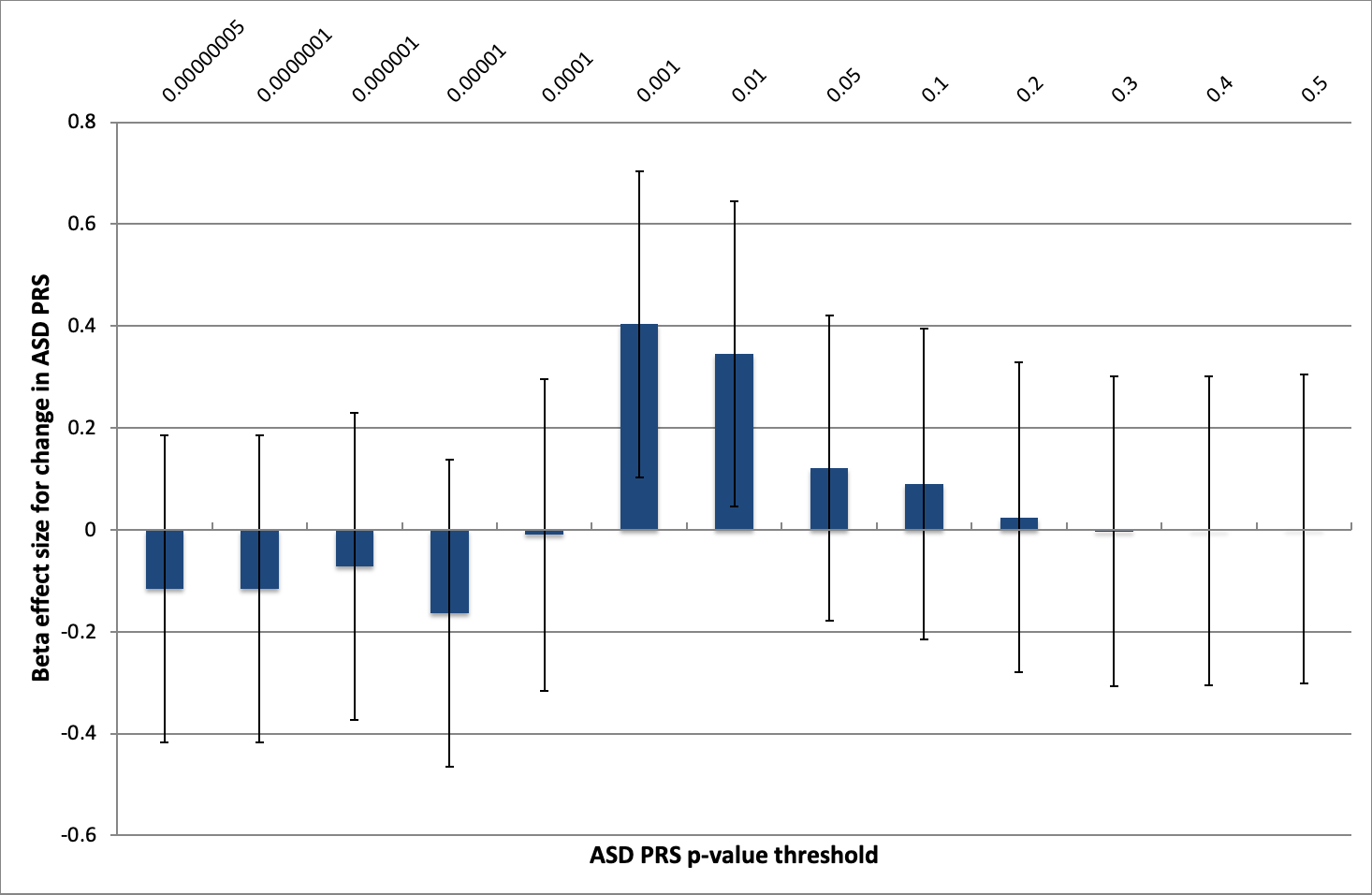

*The bars represent the effect size of the association with 95% confidence intervals shown. PRS=polygenic risk score, ASD=autism spectrum disorder, ERT=emotion recognition task, ALSPAC=Avon longitudinal study of parents and children*

**Supplementary Figure S10. Plot of associations between standardised ASD polygenic risk score and the DANVA outcome at age 8 in ALSPAC.**

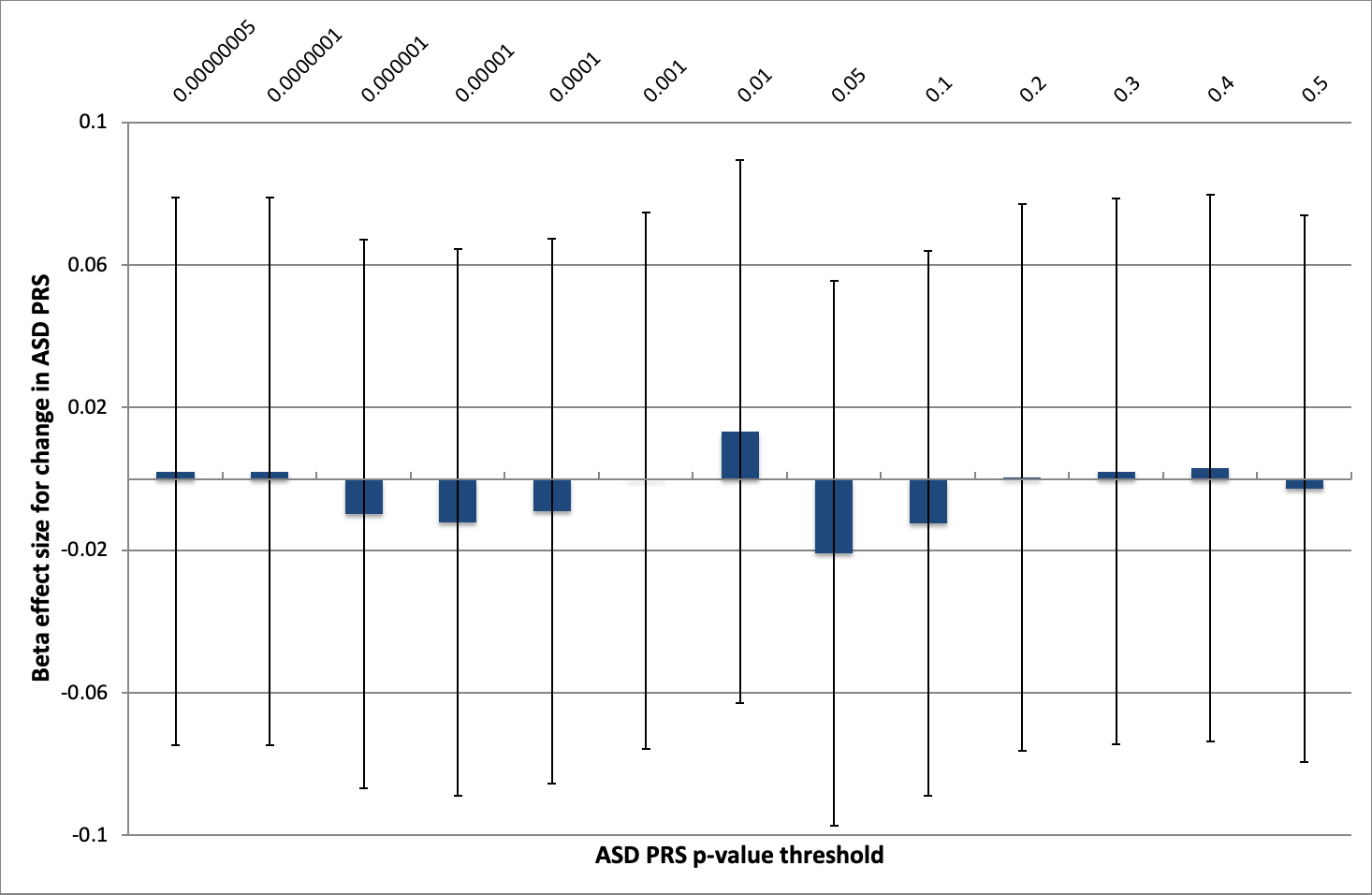

*The bars represent the effect size of the association with 95% confidence intervals shown. PRS=polygenic risk score, ASD=autism spectrum disorder, DANVA=* *Diagnostic Analysis of Nonverbal Accuracy, ALSPAC=Avon longitudinal study of parents and children*
